# REM sleep rhythm impairment in people with PTSD

**DOI:** 10.1101/2025.07.02.25330609

**Authors:** Haruka Sekiba, Sayuri Hirohata, Arinobu Hori, Yosuke Suga, Yuko Toshishige, Pablo Vergara, Minori Masaki, Morie Tominaga, Masashi Yanagisawa, Yoshiharu Kim, Masanori Sakaguchi

**Author notes:** Corresponding author (Masanori Sakaguchi, International Institute for Integrative Sleep Medicine (WPI-IIIS), University of Tsukuba, Ibaraki, 305-8587, Japan,).

## Abstract

**Study objectives:** Sleep disturbances are reported by approximately 90% of people with post-traumatic stress disorder (PTSD), yet objective characterizations of their sleep remain inconsistent across studies. This inconsistency likely stems from the limitations of standard, single-night, laboratory-based polysomnography (PSG), which often fails to capture naturalistic sleep patterns in hypervigilant individuals. To address this critical gap, our objective was to characterize the architecture of naturalistic sleep using multi-night, in-home PSG. A more precise understanding of these sleep abnormalities is crucial for advancing our knowledge of PTSD pathophysiology and is expected to provide a foundation for developing novel, targeted therapeutic strategies.

**Methods:** In-home sleep recordings using only five electrodes and a portable PSG device were obtained from 29 people with PTSD and 29 matched healthy control individuals across multiple nights.

**Results:** Although total sleep time was similar between people with PTSD and matched healthy control individuals, people with PTSD exhibited disruptions in several rapid eye movement (REM) sleep parameters, including prolonged latency to REM sleep, shorter REM sleep episodes, fragmentation of REM sleep, and weaker REM sleep rhythmicity.

**Conclusion:** These results offer novel insights into the sleep pathophysiology of PTSD by identifying REM sleep as a core domain of disruption. These findings also underscore the urgent need for further research into how REM sleep disturbances contribute to PTSD symptoms and highlight the importance of ecologically valid sleep assessments and REM sleep-targeted therapeutic strategies for more effective treatment of PTSD.

**Clinical trial registration:** Registry: jRCT (Japan Registry of Clinical Trials): https://jrct.mhlw.go.jp/

Name: An Observational Study to Acquire Patient EEG Data for the Development of a Device Enabling Acoustic Exposure Therapy during Sleep for Post-Traumatic Stress Disorder (PTSD).

Registration number: jRCT1032220029

**Statement of significance:** Posttraumatic stress disorder (PTSD) is commonly associated with sleep-related disturbances, particularly trauma-related nightmares. Using multi-night in-home polysomnography, this study characterizes people with PTSD’ naturalistic sleep patterns. While total sleep time was comparable between people with PTSD and healthy control individuals, people with PTSD exhibited disruptions in several aspects of rapid eye movement (REM) sleep, including weaker REM sleep rhythmicity across the night. These findings offer novel insights into the sleep pathophysiology of PTSD by identifying REM sleep as a core domain of disruption and further highlight the importance of ecologically valid sleep assessments and REM sleep-targeted therapeutic strategies for more effective treatment of PTSD.

## Introduction

Sleep disturbances are highly prevalent among individuals with psychiatric conditions, including depression, anxiety disorders, and cognitive impairment [1]. These disturbances can be not only symptoms but also contributing factors that exacerbate the underlying psychopathology [2]. Among these conditions, post-traumatic stress disorder (PTSD), which can arise in the aftermath of life-threatening or severely traumatic events, is particularly associated with sleep disturbances. The Diagnostic and Statistical Manual of Mental Disorders, Fifth Edition, Text Revision (DSM-5-TR), includes sleep-related symptoms within its diagnostic criteria for PTSD, notably under Criterion B (intrusive symptoms) and Criterion E (alterations in arousal and reactivity). Consistent with these diagnostic features, more than 90% of people with PTSD report sleep-related problems, and over half experience recurrent nightmares [3, 4].

Although sleep disturbances are widely prevalent in PTSD, objective sleep measurements have produced inconsistent findings, complicating efforts to characterize the underlying sleep pathology. For example, whereas some studies report that total sleep time in people with PTSD is comparable to that in healthy controls [5], others document significant reductions [1, 6]. Similarly, findings regarding rapid eye movement (REM) sleep—the stage most closely associated with dreaming—are mixed. Some studies reported shortened REM sleep latency and increased REM density, referring to the frequency of rapid eye movements [1, 5], whereas others describe the fragmentation of REM sleep, including shortened and increased REM sleep episodes and frequent transitions to wakefulness [7, 8].

This variability may stem from a range of contributing factors, such as age, sex, comorbid conditions, medication use, and differences in recording environments or devices [6]. Polysomnography (PSG), the gold standard for objective sleep assessment, enables precise identification of sleep stages through electroencephalographic (EEG) recordings. However, standard PSG typically requires an overnight stay in a hospital or laboratory and the attachment of numerous body sensors, which can be physically and psychologically intrusive. Moreover, PSG is susceptible to the so-called “the first-night effect,” in which sleep architecture during the initial night of laboratory monitoring is often altered—characterized by increased wakefulness and reduced sleep depth—compared with habitual sleep patterns [9]. This effect underscores the influence of unfamiliar environments on sleep quality. In people with PTSD, such unfamiliar settings may provoke heightened vigilance, further amplifying the first-night effect. As a result, conventional PSG conducted in laboratory settings may fail to capture the representative sleep architecture in people with PTSD.

In this study, we investigated sleep architecture in people with PTSD across multiple nights using portable PSG in their home environment. This approach was designed to accurately characterize habitual sleep patterns while enabling precise determination of sleep stages. By conducting recordings in a familiar and comfortable setting, we aimed to obtain ecologically valid data that better represents typical sleep architecture in people with PTSD.

## Methods

### Ethical Considerations

This study was conducted in accordance with the principles of the Declaration of Helsinki. The study protocol was reviewed and approved by the Research Ethics Committee of the Faculty of Engineering, Information and Systems, University of Tsukuba (Approval No. 2023R728). All participants provided written informed consent prior to their participation in the study. The study method and results are reported following the Strengthening the Reporting of Observational Studies in Epidemiology (STROBE) Statement.

### Sleep recordings from people with PTSD

A total of 41 people with PTSD were recruited through referrals from eight physicians across Japan. All patients were native Japanese speakers aged between 20 and 65 years and were able to provide written informed consent after receiving a detailed explanation of the study procedures. Eligibility for participation was determined using the Posttraumatic Diagnostic Scale (PDS-IV), a self-report questionnaire designed to diagnose and assess PTSD symptoms based on DSM-IV criteria, including intrusion, avoidance, negative alterations in cognition and mood, and alterations in arousal and reactivity. The PDS-IV was used due to the absence of a validated Japanese version of the PDS-5 at the time of study initiation. To ensure that patients still met PTSD diagnostic criteria at the time of sleep recordings, the interval between PDS-IV completion and the start of PSG recordings was required to be ≤6 weeks. Exclusion criteria included severe sleep disorders known to interfere with PSG recordings, such as severe obstructive sleep apnea (OSA), restless legs syndrome (RLS), and REM sleep behavior disorder (RBD). There were no restrictions on the use of psychiatric or sleep-related medications. Patients for whom a matched healthy controls could not be identified were also excluded. After applying these criteria, 29 people with PTSD were included in the final analysis (Figure 1, Table 1).

**Figure 1.**
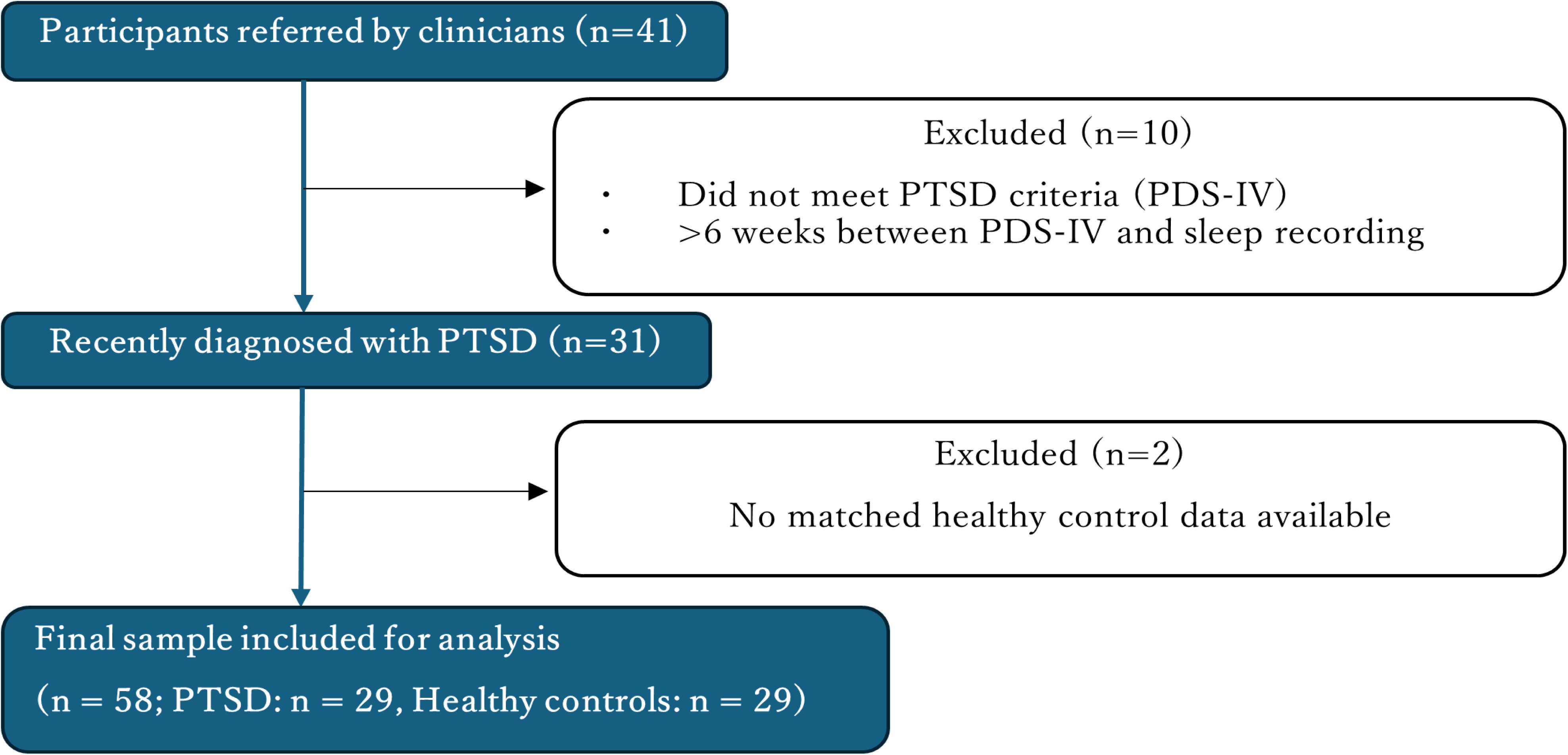
Participant recruitment flow.

**Table 1.** Participant demographic information. The absolute standardized difference in variables for propensity score estimation was used to assess the match balance between people with PTSD and healthy control groups. A difference of ≤ 0.10 for all parameters indicates an acceptable match balance. BMI, body mass index; PTSD, post-traumatic stress disorder.

|  | Healthy | PTSD | Absolute standardized difference |
| --- | --- | --- | --- |
| Female, n (%) | 22 (76%) | 21 (72%) | 0.079 |
| Age, mean ± SD [years] | 45 ± 9.1 | 43 ± 13 | 0.10 |
| BMI, mean ± SD [kg/m<sup>2</sup>] | 24 ± 4.1 | 24 ± 4.3 | 0.081 |

Sleep recordings were made using the portable Insomnograf K2 (S’UIMIN Inc., Tokyo, Japan) PSG device. The quality of sleep recording by the Insomnograf K2 is comparable to that of a typical PSG device, achieving an 86.9% concordance rate and kappa coefficient of 0.80 (Seol et al., 2022; Masaki et al., 2025), which is higher than the kappa coefficient of 0.76 for a typical PSG device (Lee et al. 2022). To ensure correct and standardized electrode placement, participants were instructed to self-apply a set of five wet, soft-gel electrodes pre-positioned on a single flexible harness. This design fixed the electrodes to their designated locations on the forehead and behind both ears. Detailed written and video instructions were provided to guide the application process. Furthermore, the device performed an automated impedance check to alert the user to inadequate electrode contact before the recording began. Any nightly recordings with signal quality deemed insufficient for reliable analysis (e.g., due to baseline drift or persistent artifacts) were excluded from the final analysis. Recording time was not pre-specified, and participants recorded their sleep at home by themselves. Each participant recorded ∼3-7 nights of sleep (mean ± standard deviation, people with PTSD: 5.1 ± 0.73, healthy controls: 4.4 ± 1.1).

From the five electrodes placed on the frontal and occipital regions, the Insomnograf generates five EEG derivations (Fp1–M2, Fp2–M1, Fp1–average M, Fp2–average M, and Fp1–Fp2). Sleep was staged in 30-second epochs by trained sleep technologists according to the American Academy of Sleep Medicine (AASM) manual (Version 2.6). The stages were classified as wakefulness after sleep onset (WASO), rapid eye movement (REM) sleep, and non-REM (NREM) sleep. NREM sleep was further categorized by depth into stage N1 (transitional sleep), stage N2 (light sleep), and stage N3. Stage N3 is characterized by high-amplitude slow-waves and is also referred to as slow-wave sleep (SWS). Additionally, arousals were scored and tagged separately in accordance with AASM scoring rules, and an arousal index was calculated as the number of arousals per hour of sleep.

### Sleep recordings from healthy controls

Healthy controls were identified from a database of healthy subjects from S’UIMIN Inc. Participants were selected from 224 people who had their EEG measured between October 2021 and December 2023 with the same equipment as that used by people with PTSD, whose EEG had been recorded between May 2022 and June 2023, and who agreed to participate in the study. As age and sex are known confounds in sleep studies (Richards et al., 2017; Boulos et al., 2019) and body-mass index (BMI) may mediate the impact of PTSD on sleep given reports that PTSD is associated with increased BMI (Suliman et al., 2016) and BMI is associated with sleep duration (Grandner et al., 2015), propensity scores were determined using logistic regression models with age, sex, and BMI covariates as independent variables. The absolute standardized difference of variables for propensity score estimation was used to assess the match balance between people with PTSD and healthy controls. The absolute standardized difference was ≤0.10 for every covariate, reflecting an acceptable match balance between groups (Table 1). The included 29 healthy controls were confirmed to have no history of active treatment for sleep-related disorders.

### Sleep recording inclusion criteria

One healthy control with only one night’s recording was excluded from sleep regularity analysis. Nights with no target sleep stage episodes were excluded from sleep latency, episode parameters, and fragmentation analyses. For all rhythmicity analyses, nights with no target sleep stages were excluded too. In addition, some individual average data that did not meet the following criteria were excluded from the rhythmicity analyses to avoid underestimating the rhythmicity of peaks. For REM sleep rhythmicity, individual average data from one healthy control and two people with PTSD were excluded because they ended within 183 min after the onset of the first REM sleep episode, coinciding with the timing of the second REM sleep autocorrelation peak in healthy controls (Figure S5a). For N3 rhythmicity, individual average data from two healthy controls and seven people with PTSD were excluded due to having <4.4 N3 episodes per night on average (Figure S5b). This threshold was set at the first quartile of the distribution of the average number of N3 episodes per night in people with PTSD to ensure sufficient data availability. For N2 rhythmicity, individual average data from one healthy control and one people with PTSD were excluded because they ended within 203.5 min after the onset of the first N2 episode, coinciding with the timing of the second N2 autocorrelation peak in healthy controls (Figure S5c). Although the onset of rhythmicity analyses was aligned to the first appearance of the target sleep stage across nights for each participant, the time at which sleep ended (i.e., final awakening) varied from night to night, resulting in differences in the duration of each recording. Consequently, when analyzing data from a common starting point, the number of available recordings gradually decreased over time, as nights with earlier sleep termination dropped out of the analysis. To account for this, individual rhythmicity was evaluated up to the time point at which ≥50% of the individual recordings remained. For instance, in participants with five nights of data, analysis was carried out until the point at which at least three nights still provided usable data. In a separate rhythmicity analysis focused on REM sleep episode interval variability, we fitted the distribution of all interval durations with a two-Gaussian mixture model to identify and exclude transient interruptions. The intersection point of the two fitted components, estimated at 17.1 minutes (Figure S6), was used as a data-driven threshold for interval classification. Intervals shorter than this threshold were excluded from the analysis. Data from 108 nights in healthy controls and 76 nights in people with PTSD were included.

### Sleep analyses

A bias-corrected and accelerated bootstrap method employing the bootci function (1,000 iterations) in MATLAB R2024a (MathWorks, Natick, MA, USA) was used to compute the 95% confidence intervals (2.5th to 97.5th percentiles) for variability in REM sleep latency and midsleep time. For rhythmicity analyses, data shuffling was performed using the randperm function (1,000 iterations) to generate a random distribution of sleep episodes. Autocorrelation was computed using the xcov function with the ‘coeff’ option, providing normalized covariance values.

For REM sleep episode interval variability analysis, a two-Gaussian mixture model was fitted to the distribution of interval durations using the MultiPeakFit 2 package in Igor Pro version 9 (WaveMetrics, Lake Oswego, OR, USA), with initial parameters for the two components manually specified to identify a threshold for classifying intervals.

Statistical analyses were conducted using GraphPad Prism version 10 (GraphPad Software, San Diego, CA, USA). Data were assessed for normality using the Shapiro Wilk test. The F-test was conducted to assess the equality of variances, and the Kolmogorov–Smirnov test was used to compare the overall distribution shapes. For normally distributed data, differences between groups were analyzed using the two-tailed unpaired t-test, with Welch’s correction if standard deviation differed between groups. Non-parametric data were analyzed using the Mann-Whitney test. Differences were considered statistically significant when *p* < 0.05.

## Results

Nightly sleep habits (Figure 2a) were examined using a sleep regularity index, which measures the consistency of sleep-wake states at the same time across recordings within participants. This measure is particularly suitable for small recording samples (<7 nights) and reflects variability in both sleep timing and duration (Fischer et al., 2021). People with PTSD had a significantly lower sleep regularity index than healthy controls (Figure 2b). However, the variation in midsleep time (midpoint between sleep onset and final awakening) was similar between groups (Figure 2c). This suggests that people with PTSD exhibit more irregular sleep habits across nights characterized by greater within-individual variability than between-individual variability.

**Figure 2.**
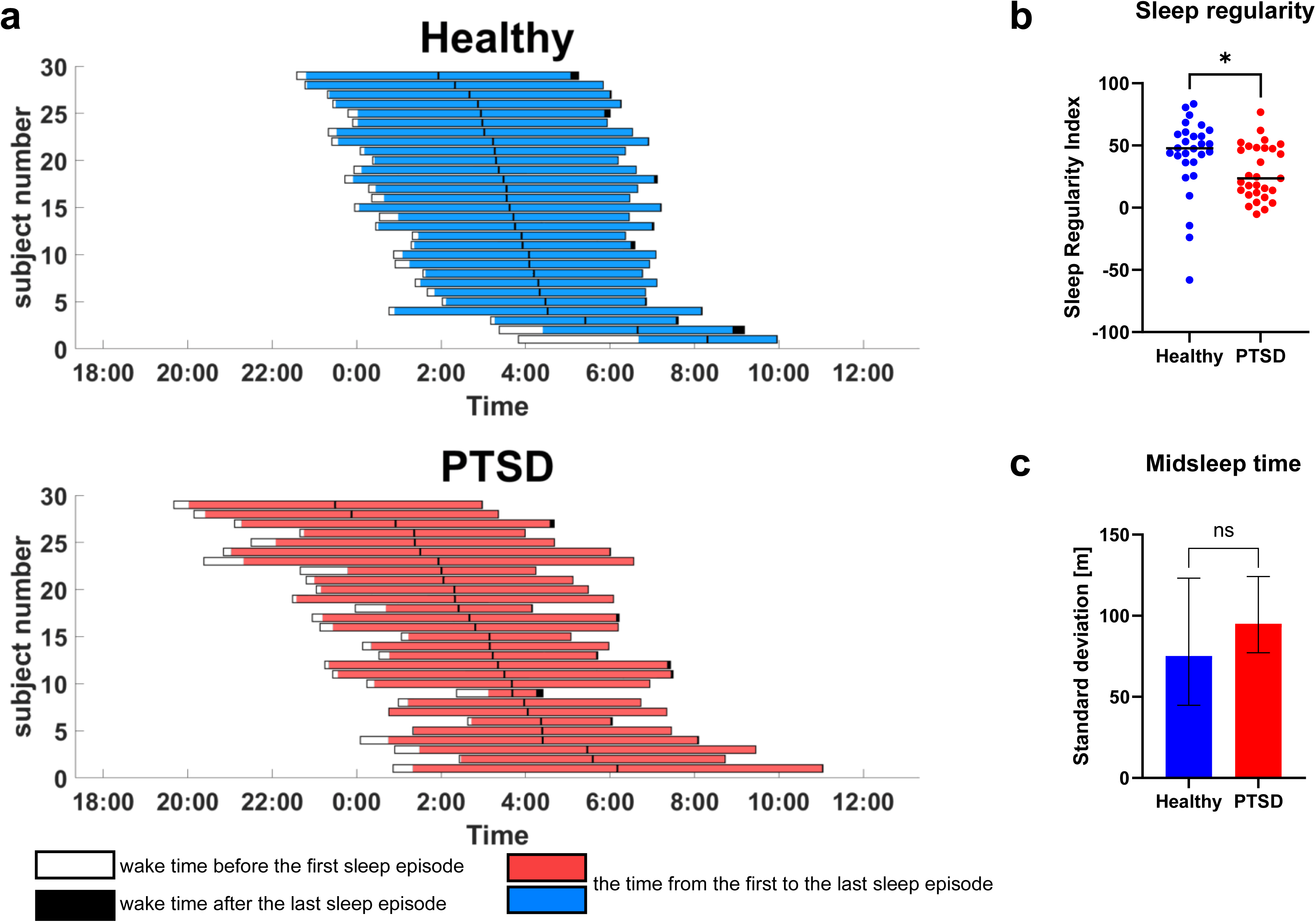
Sleep regularity. (a) Average schedule across several sleep recordings for individual healthy controls and people with PTSD. The central black line represents the midsleep time. (b) Sleep regularity index for participants with more than two nights of sleep recordings. People with PTSD exhibited less sleep regularity than healthy controls (*p* = 0.014). (c) Standard deviation of midsleep time within groups. Variation in midsleep time was similar between groups (*p* = 0.22). Error bars represent the 95% CI, calculated using bootstrap resampling, Healthy [44.78, 123.21]; PTSD [77.13, 124.21]. \**p* < 0.05. CI, confidence interval; ns, not significant; PTSD, post-traumatic stress disorder.

Total sleep time was similar between people with PTSD and healthy controls (Figure 3a), but sleep latency was significantly prolonged in people with PTSD (Figure 3b). In accordance with these observations, sleep efficiency (percentage of time asleep while in bed) was slightly but significantly lower in people with PTSD (Figure 3c). Moreover, people with PTSD exhibited a significantly higher frequency of arousals compared to healthy controls (Figure 3d). Together, these observations suggest that people with PTSD have difficulties not in the quantity of sleep, but in initiating and maintaining it.

**Figure 3.**
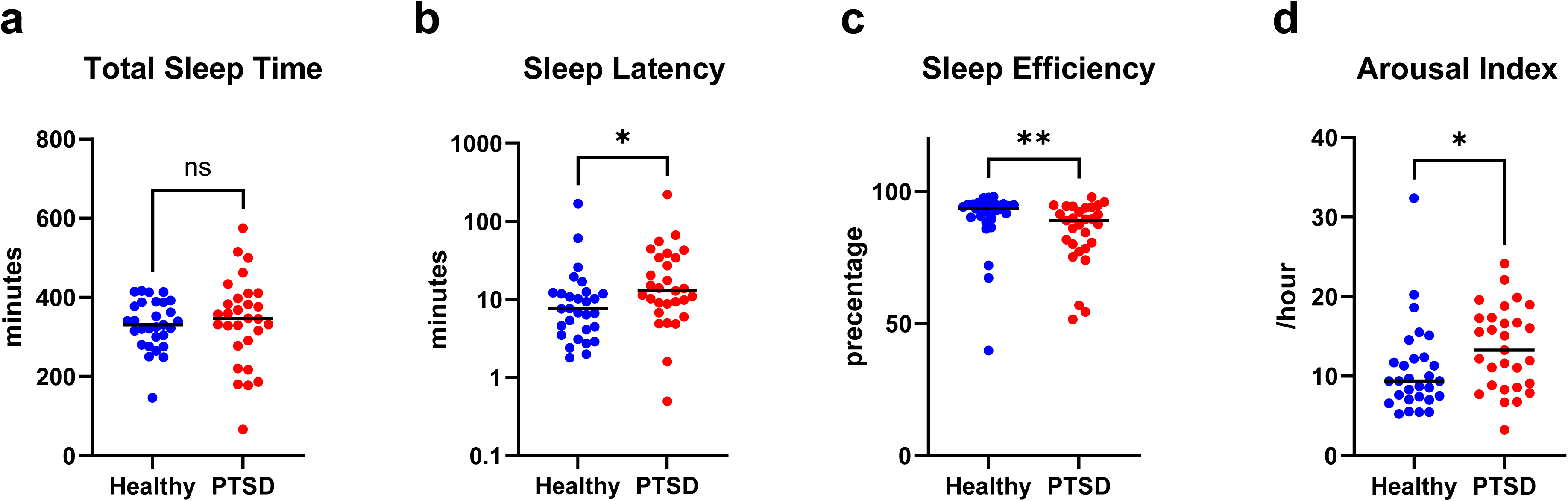
Total sleep time, latency, efficiency, and Arousal Index. Each data point represents the average across several nights for each participant. Y axis in (b) is plotted on a log₁₀ scale. (a) Total sleep time was similar between groups (*p* = 0.70), whereas (b) sleep latency was prolonged (*p* = 0.018), (c) sleep efficiency was lower (*p* = 0.0096), and (d) Arousal Index was higher (*p* = 0.020) in people with PTSD than in healthy controls. \**p* < 0.05. \*\**p* < 0.01. ns, not significant; PTSD, post-traumatic stress disorder.

Considering each sleep stage in detail, people with PTSD exhibited significantly more N2 and less REM sleep than healthy controls, with similar amounts of N1 and N3 between groups (Figure 4a). However, many people with PTSD showed almost no N3, which was underscored by a significant difference in the distribution of N3 durations between groups (Figure S1). People with PTSD also showed significantly prolonged latency to REM sleep (Figure 4b) and significantly shorter REM sleep episodes (Figure 4c) than healthy controls. In contrast to these abnormalities in REM sleep, there were generally no further differences between groups in non-REM sleep parameters (Figure 4b-d). Although latency to N1 was significantly different between groups (Figure 4b), both groups had a median of 0 and a small interquartile range (healthy controls: 0, people with PTSD: 0.40), suggesting that the difference was driven by a few outliers in people with PTSD. Moreover, the REM sleep fragmentation index (number of transitions from REM sleep to WASO per hour of REM sleep) was significantly higher in people with PTSD than in healthy controls (Figure 4e). This indicates that REM sleep episodes were frequently disrupted by wakefulness, consistent with significantly more WASO episodes per night in people with PTSD (Figure 4d). Together, these results suggest that people with PTSD struggle both to initiate and maintain REM sleep, leading to less REM sleep overall.

**Figure 4.**
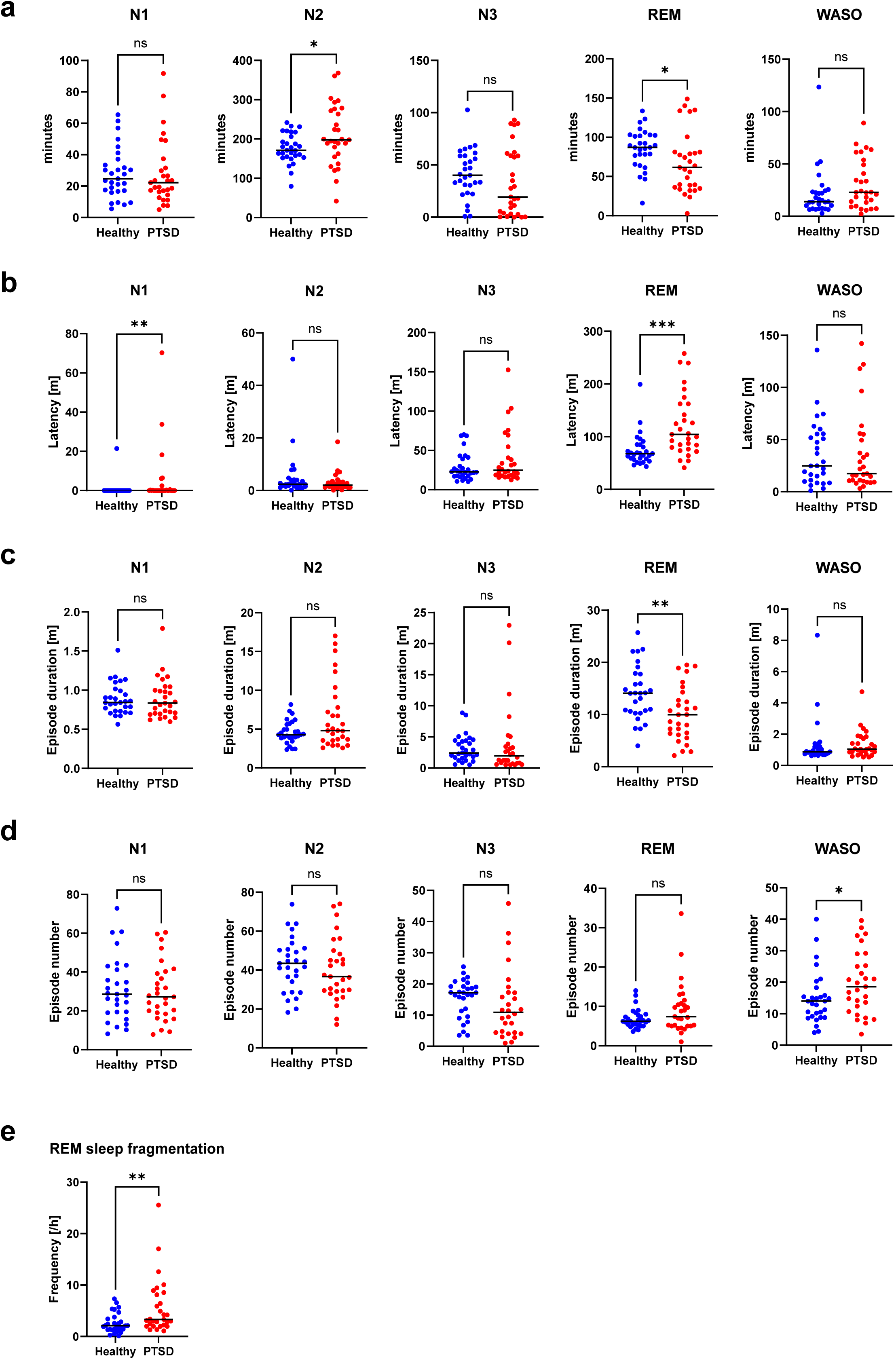
Sleep stage amount, latency, episode duration and number, and fragmentation. Each data point represents the average across several nights for each participant. (b-e) reflect only nights when the target sleep stage was present. (a) People with PTSD showed more N2 sleep and less REM sleep than healthy controls (N1: *p* = 0.58, N2: *p* = 0.042, N3: *p* = 0.14, REM: *p* = 0.045, WASO: *p* = 0.073). (b) People with PTSD showed prolonged latency from sleep onset to the first N1 and REM episodes compared with healthy controls (N1: *p* = 0.0045, N2: *p* = 0.42, N3: *p* = 0.34, REM: *p* < 0.001, WASO: *p* = 0.64). (c, d) The duration and number of sleep stage episodes were similar between groups except for a shorter REM episode duration and larger WASO episode number ((c) N1: *p* = 0.72, N2: *p* = 0.20, N3: *p* = 0.29, REM: *p* = 0.0042, WASO: *p* = 0.33; (d) N1: *p* = 0.75, N2: *p* = 0.48, N3: *p* = 0.051, REM: *p* = 0.23, WASO: *p* = 0.049). (e) REM sleep fragmentation was higher in people with PTSD than in healthy controls (*p* = 0.0033). \**p* < 0.05, \*\**p* < 0.01, \*\*\**p* < 0.001. ns, not significant; PTSD, post-traumatic stress disorder; REM, rapid eye movement; WASO, wakefulness after sleep onset.

Next, we analyzed sleep stage cycles over the course of a night. Strikingly, two peaks observed in healthy controls—a “first REM sleep peak” occurring approximately 75 min after sleep onset and a “second N3 peak” occurring around 110 min after sleep onset—were not observed in people with PTSD (Figure 5, Figure S2).

**Figure 5.**
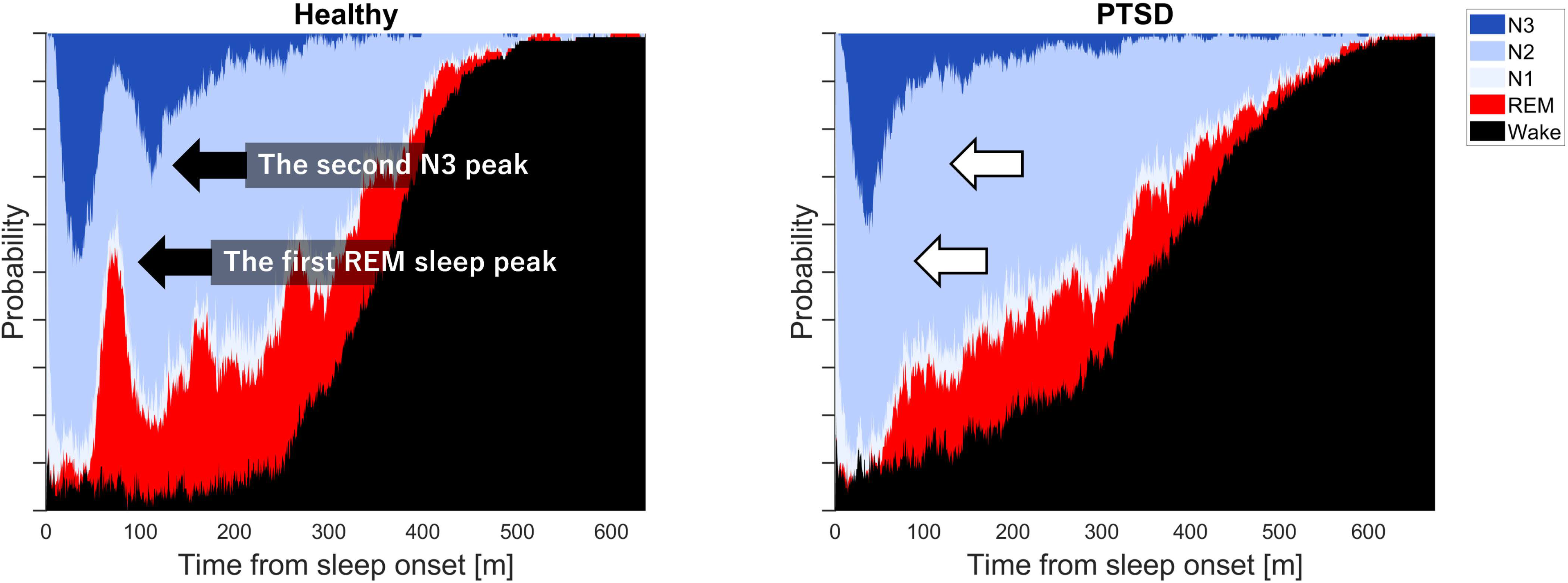
Sleep stage peaks. Stacked area chart depicting all sleep recordings (Healthy: 130 nights; PTSD: 149 nights). The x-axis is aligned to 0, representing the onset of the first sleep episode. In healthy controls, the first REM sleep peak occurred around 75 minutes and the second N3 peak around 110 minutes. These peaks were absent in people with PTSD. PTSD, post-traumatic stress disorder; REM, rapid eye movement.

As people with PTSD showed significantly more variability in REM sleep latency than healthy controls (Figure 6b), it is possible that individual REM sleep peaks in people with PTSD were averaged out in Figure 6a. However, even when the timing of the first REM sleep episode onset was aligned across individuals, people with PTSD still lacked clear REM sleep peaks, unlike healthy controls (Figure 6c). To directly investigate REM sleep rhythmicity, we first generated “shuffled data” by randomizing the occurrence of REM sleep episodes and intervals between episodes within nights. We then calculated the autocorrelation of REM sleep probability and defined “rhythmicity” as the root mean square deviation from the shuffled data (Figure 6d). This rhythmicity analysis for each participant revealed that people with PTSD exhibited significantly weaker REM sleep rhythmicity than healthy controls (Figure 6e). Given that these analyses are based on group-or individual-averaged data, it still remains possible that differences in rhythmicity across nights among people with PTSD were averaged out. To address this, we examined the standard deviation of the intervals between REM sleep episodes within nights as an additional rhythmicity parameter. Consistently, we observed significantly weaker rhythmicity in people with PTSD than in healthy controls, as evidenced by a wider standard deviation of REM sleep episode intervals (Figure 6e).

**Figure 6.**
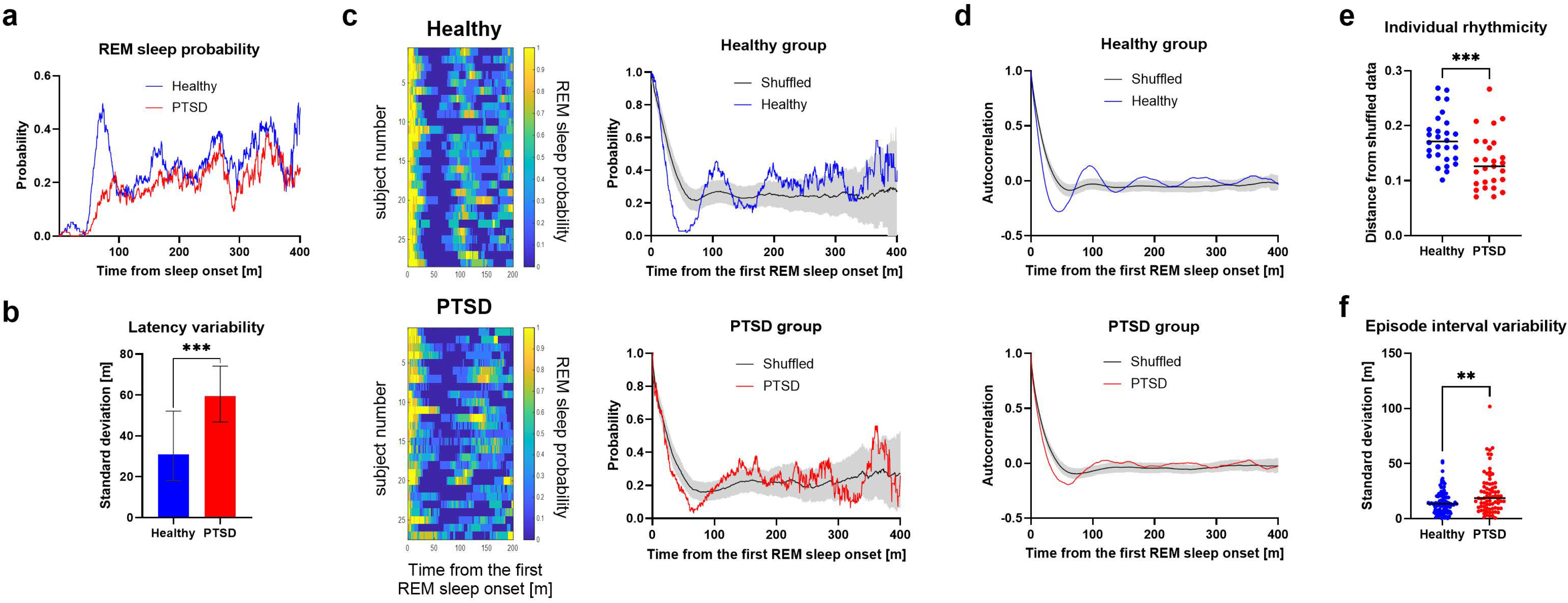
REM sleep rhythmicity. (a, b) Group-average REM sleep probability and group-standard deviation of REM sleep latency calculated from individual averages across multiple nights. (a) The x-axis is aligned to 0, representing the onset of the first sleep episode. People with PTSD lacked the first REM sleep peak observed in healthy controls. (b) Nights with no REM sleep were excluded. People with PTSD showed greater variability in REM sleep latency than healthy controls (*p* < 0.001). Error bars represent the 95% CI, calculated using bootstrap resampling, Healthy [18.12, 52.13]; PTSD [46.78, 74.10]. (c-f) Nights with no REM sleep or individual average data ending within 183 min after the onset of the first REM sleep episode were excluded. (c, d) Pattern of REM sleep episode occurrence after the first episode. Heatmap showing individual-average probability (c left), group-average probability (c right), and its autocorrelation (d). The x-axes are aligned to 0, representing the onset of the first REM sleep episode. The shuffled dataset consists of 1000 group averages, each derived from individual averages across multiple nights, with REM sleep episodes and their intervals shuffled within nights. The black line is the mean of the shuffled data, and shaded areas represent the 95% CI derived from shuffled dataset distributions. People with PTSD still lacked the first REM sleep peak. (e) Each data point represents the root mean square deviation of individual average autocorrelation from their respective shuffled data, evaluated up to the time point at which ≥50% of the individual recordings remained. People with PTSD showed weaker REM sleep rhythmicity than healthy controls (*p* < 0.001). (f) Each data point represents the standard deviation of intervals (>17.1 min) between REM sleep episodes within nights. REM sleep episodes occurred more randomly in people with PTSD than in healthy controls (*p* = 0.0019). \*\**p* < 0.01, \*\*\**p* < 0.001. CI, confidence interval; PTSD, post-traumatic stress disorder; REM rapid eye movement.

The same rhythmicity analysis for N3 (Figure 7). We noticed that there is the first peak in both groups around 70 min after sleep onset. However, even after aligning the timing of the first N3 episode onset, the second N3 peak that was present in healthy controls remained absent in people with PTSD (Figure 7b). Accordingly, people with PTSD exhibited significantly weaker N3 rhythmicity than healthy controls (Figure 7c, d). Due to the small number of N3 episodes, episode interval analysis for N3 was not conducted. By contrast, N2 rhythmicity was similar between groups (Figure S3), and there were no rhythmic patterns in N1 or WASO in either group (Figure S4).

**Figure 7.**
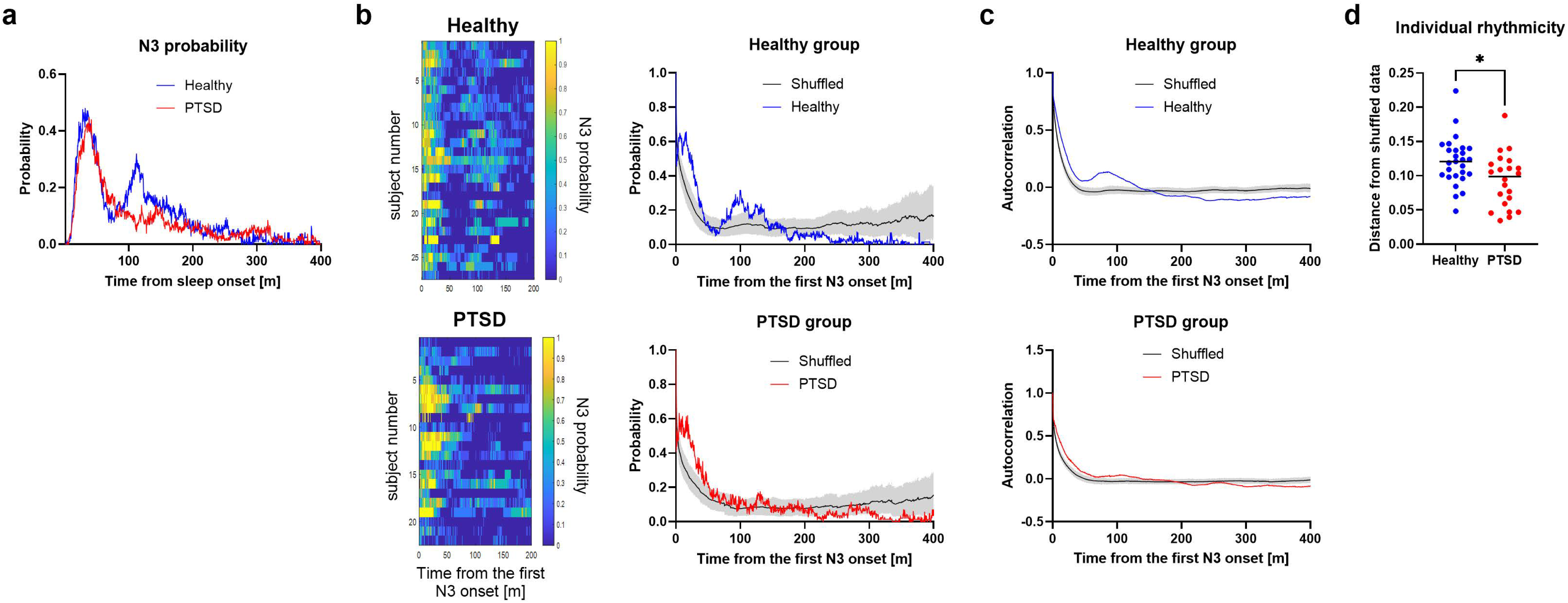
N3 rhythmicity. (a) Group-average N3 probability calculated from individual averages across multiple nights. The x-axis is aligned to 0, representing the onset of the first sleep episode. People with PTSD lacked the second N3 peak observed in healthy controls. (b-d) Nights with no N3 or individual average data with <4.4 N3 episodes per night on average were excluded. (b, c) Pattern of N3 episode occurrence after the first episode. Heatmap showing individual-average probability (b left), group-average probability (b right), and its autocorrelation (c). The x-axes are aligned to 0, representing the onset of the first N3 episode. The shuffled dataset consists of 1000 group averages, each derived from individual averages across multiple nights, with N3 episodes and their intervals shuffled within nights. The black line is the mean of the shuffled data, and shaded areas represent the 95% CI derived from shuffled dataset distributions. People with PTSD still lacked the second N3 peak. (d) Each data point represents the root mean square deviation of individual average autocorrelation from their respective shuffled data, evaluated up to the time point at which ≥50% of the individual recordings remained. People with PTSD showed weaker N3 rhythmicity than healthy controls (p = 0.011). \**p* < 0.05. CI, confidence interval; PTSD, post-traumatic stress disorder.

## Discussion

As PTSD is commonly associated with sleep disturbances, we sought to further understand people with PTSD’ naturalistic sleep architecture using multi-night in-home PSG. We found that people with PTSD showed irregular sleep habits, prolonged sleep latency, and higher arousal index compared with healthy controls. Sleep stage-specific analysis revealed rather normal non-REM sleep parameters in people with PTSD, except for a near absence of N3 in some patients. By contrast, a variety of abnormalities in REM sleep parameters were observed in people with PTSD, including delayed onset, reduced duration, and fragmentation. Furthermore, we found a disruption in REM sleep rhythmicity in people with PTSD.

Previous studies examining one-or two-night PSG recordings, mostly performed in non-habitual environments (e.g., hospital or laboratory) report that people with PTSD have less total sleep time, and N3 although findings vary across studies [1, 5, 6]. However, no such finding was observed in our study, which examined approximately three to seven nights of recordings in a habitual environment (i.e., home). Notably, previous studies with one-night PSG recording performed at home report that sleep macro-architecture did not differ significantly between individuals with PTSD and trauma-exposed individuals without PTSD [18, 19]. These mixed findings, including those from our study, suggest that the recording environment and the number of nights may influence sleep characteristics in people with PTSD. Indeed, prior studies using actigraphy across multiple-night recordings in habitual environments reported reduced sleep regularity in people with PTSD [20–22], consistent with the current results. Overall, these findings underscore the importance of multiple-night recordings in a habitual environment for the analysis of regular sleep patterns in people with PTSD, who commonly experience hyper-vigilance in unfamiliar environments.

REM sleep, originally identified as a state characterized by vivid dreaming [23], contains disproportionately negative emotional content in young individuals with depression and anxiety [24]. REM sleep disturbances have also long been recognized as key features of PTSD [25, 26]. Previous studies using PSG mostly performed in non-habitual environments report shortened REM sleep latency and increased REM density in people with PTSD [1, 5]. By contrast, we observed prolonged REM sleep latency and shorter REM sleep episodes in people with PTSD. We also observed that people with PTSD exhibit heightened arousal, particularly characterized by frequent awakenings during REM sleep (i.e., REM sleep fragmentation). Such REM sleep fragmentation is associated with the severity of early symptoms [27, 28] and nightmares [29] in people with PTSD.

REM sleep is thought to play a crucial role in emotional adaptation [27, 30]. Disruption of REM sleep impairs fear extinction in both humans [31, 32] and rodents [33, 34]. Indeed, a key feature of PTSD is resistance to fear (i.e., traumatic) memory extinction, which has been linked to disturbed REM sleep [35] and involves amygdala hyperactivity and, hippocampal and ventral-medial prefrontal cortex hypoactivity [36, 37]. These findings suggest that REM sleep dysfunction may underlie the core pathophysiology of PTSD.

The key finding of this study is that people with PTSD show weaker REM sleep rhythmicity that appears to affect N3 rhythmicity. Indeed, REM sleep plays an important role in the overall sleep cycle, as the number and length of sleep cycles are more strongly associated with REM-related than NREM-related sleep parameters [38, 39]. In both rodents [40–42] and humans [43], the duration of an REM sleep episode is positively correlated with the interval until the next REM sleep episode as well as the amount of NREM sleep occurring during that interval. As a possible mechanistic link, a rodent study shows that slow-wave activity, a hallmark of N3, is modulated by prior REM sleep and is reduced by REM sleep deprivation [44]. Together, these findings suggest that impaired REM sleep rhythmicity may cascade into irregularity and dysfunction in NREM sleep, especially N3, in people with PTSD.

Sleep is influenced by numerous factors, including comorbid illnesses and medications, which were not accounted for in this study. Notably, selective serotonin reuptake inhibitors, the first-line drug for PTSD, prolongs REM sleep latency and decreases REM sleep duration [45–47], consistent with the sleep characteristics of people with PTSD observed in our study. Despite this limitation, our study highlights the potential benefits of treating sleep disturbances in PTSD beyond simply increasing sleep quantity. For example, cognitive behavioral therapy for insomnia (CBT-I), especially sleep restriction and stimulus control, may reduce sleep latency and improve sleep efficiency [48]. Indeed, it is recommended as the first-line treatment for PTSD-related sleep issues in the VA/DoD Clinical Practice Guideline (CPG), as it alleviates both insomnia and PTSD symptoms [49, 50].

Although specifically targeting REM sleep is challenging, prazosin, an alpha-1 adrenergic antagonist that reduces noradrenergic activity, has emerged as a potential treatment for REM sleep-related abnormalities such as nightmares. Although its effects are partial, prazosin is associated with improvements in both sleep and PTSD symptoms [51–53]. Some studies report that prazosin increases REM sleep duration and episode length, contributing to broader symptom relief [54]. Further research is warranted to better understand how improving REM sleep quality may contribute to alleviation of PTSD symptoms.

Overall, the present study suggests that multi-night recordings using PSG at home can provide new perspectives on sleep abnormalities in people with PTSD. In addition to traditional sleep assessments, focusing on REM sleep abnormalities, particularly its rhythmicity, may be crucial for advancing our understanding of PTSD and developing more effective treatments.

## Data Availability

All data produced in the present study are available as described in the manuscript and supplementary material. Raw EEG data are available upon reasonable request to the corresponding author, with permission from S′UIMIN Inc.

https://github.com/seki-haru/PTSD_Sleep_Analysis.git

https://data.mendeley.com/preview/5j4y36cb5c?a=17d52a35-d4d0-486d-b0a1-57a905709669

## Acknowledgments

We thank Drs. M. Oe, S. Lin, M. Maeda, and T. Seki for recruiting patients; Drs. K. Kuriyama, A. Kawamura, K. Ino, and T. Kambayashi for valuable suggestions; I. Park and S. Nakamura for technical support; and I. Kimura, M. Sakurai, and I. Sekiguchi for administrative support. This work was partially supported by the Japan Agency for Medical Research and Development (AMED) (JP21zf0127005, JP23wm0525003, JP21km0908001), Japan Society for the Promotion of Science (JSPS) (JP24H00894, JP23H02784, JP22H00469, JP16H06280), and Takeda Science Foundation to MS; JSPS (JP23KJ0286) to MM.

## Data and Code availability

The data underlying the figures in this article are available in the article and in its online supplementary material. The raw electroencephalography (EEG) data are not publicly available as they are the property of a third party, S’UIMIN Inc. These data can be shared on reasonable request to the corresponding author with permission from S’UIMIN Inc. Custom MATLAB scripts used for sleep parameter analysis (excluding the parameters presented in Figure 3 and Figure 4a, which were reported by sleep technologists) are available on GitHub: https://github.com/seki-haru/PTSD_Sleep_Analysis.git. The repository also includes the 30-second epoch sleep stage data used in the analysis. The data supporting the plots in this study are available on Mendeley Data: https://data.mendeley.com/preview/5j4y36cb5c?a=17d52a35-d4d0-486d-b0a1-57a905709669. The participant IDs and recording IDs used in the dataset were independently assigned by S’UIMIN Inc. and cannot be used to identify individuals.

## Quantification and Statistical Analysis

Figure 2b. Healthy (n = 28) versus PTSD (n = 29), Mann–Whitney test, *U* = 252, *p* = 0.014

Figure 2c. Healthy (n = 29) versus PTSD (n = 29), F-test, *F*(28, 28) = 1.6, *p* = 0.22. 95% CI: Healthy = [44.78, 123.21]; PTSD = [77.13, 124.21]

Figure 3a. Healthy (n = 29) versus PTSD (n = 29), Two-tailed Welch’s t-tests, *t*(44) = 0.39, *p* = 0.70

Figure 3b. Healthy (n = 29) versus PTSD (n = 29), Mann–Whitney test, *U* = 269.5, *p* = 0.018

Figure 3c. Healthy (n = 29) versus PTSD (n = 29), Mann–Whitney test, *U* = 255, *p* = 0.0096

Figure 3d. Healthy (n = 29) versus PTSD (n = 29), Mann–Whitney test, *U* = 272, *p* = 0.020

Figure 4a.

N1: Healthy (n = 29) versus PTSD (n = 29), Mann–Whitney test, *U* = 384, *p* = 0.58

N2: Healthy (n = 29) versus PTSD (n = 29), Two-tailed Welch’s t-tests, *t*(41) = 2.1, *p* = 0.042 N3: Healthy (n = 29) versus PTSD (n = 29), Mann–Whitney test, *U* = 326, *p* = 0.14

REM: Healthy (n = 29) versus PTSD (n = 29), Two-tailed Welch’s t-tests, *t*(48) = 2.1, *p* = 0.045 WASO: Healthy (n = 29) versus PTSD (n = 29), Mann–Whitney test, *U* = 305, *p* = 0.073

Figure 4b-d.

N1: Healthy (n = 29) versus PTSD (n = 29), (b) Mann–Whitney test, *U* = 293, *p* = 0.0045; (c) Mann–Whitney test, *U* = 397, *p* = 0.72; (d) Mann–Whitney test, *U* = 400, *p* = 0.75

N2: Healthy (n = 29) versus PTSD (n = 29), (b) Mann–Whitney test, *U* = 368.5, *p* = 0.42; (c) Mann–Whitney test, *U* = 338, *p* = 0.20; (d) Two-tailed unpaired t-tests, *t*(56) = 0.71, *p* = 0.48 N3: Healthy (n = 29) versus PTSD (n = 28), (b) Mann–Whitney test, *U* = 345, *p* = 0.34; (c) Mann–Whitney test, *U* = 339.5, *p* = 0.29; (d) Mann–Whitney test, *U* = 284, *p* = 0.051

REM: Healthy (n = 29) versus PTSD (n = 29), (b) Mann–Whitney test, *U* = 181, *p* < 0.001; (c) Two-tailed unpaired t-tests, *t*(56) = 3.0, *p* = 0.0042; (d) Mann–Whitney test, *U* = 343, *p* = 0.23 WASO: Healthy (n = 29) versus PTSD (n = 29), (b) Mann–Whitney test, *U* = 390, *p* = 0.64; (c) Mann–Whitney test, *U* = 357, *p* = 0.33; (d) Mann–Whitney test, *U* = 294, *p* = 0.049

Figure 4e. Healthy (n = 29) versus PTSD (n = 29), Mann–Whitney test, *U* = 234, *p* = 0.0033

Figure 6b. Healthy (n = 29) versus PTSD (n = 29), F-test, *F*(28, 28) = 3.7, *p* < 0.001. 95% CI: Healthy = [18.12, 52.13]; PTSD = [46.78, 74.10]

Figure 6d. Healthy (n = 28) versus PTSD (n = 27), Mann–Whitney test, *U* = 180, *p* < 0.001

Figure 6e. Healthy (n = 108) versus PTSD (n = 76), Mann–Whitney test, *U* = 3006, *p* = 0.0019

Figure 7c. Healthy (n = 27) versus PTSD (n = 22), Two-tailed unpaired t-tests, *t*(47) = 2.6, *p* = 0.011

Figure S1. Healthy (n = 29) versus PTSD (n = 29), Kolmogorov–Smirnov test, *D* = 0.38, *p* = 0.031

Figure S3e. Healthy (n = 28) versus PTSD (n = 28), Mann–Whitney test, *U* = 377, *p* = 0.81

