## Supplementary materials for "REM sleep rhythm impairment in people with PTSD"

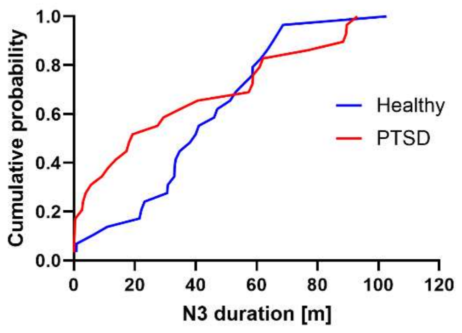


**Figure S1. Cumulative probability distribution of N3 duration.** People with PTSD had a higher proportion of shorter N3 durations ($p$ = 0.031). PTSD, post-traumatic stress disorder.

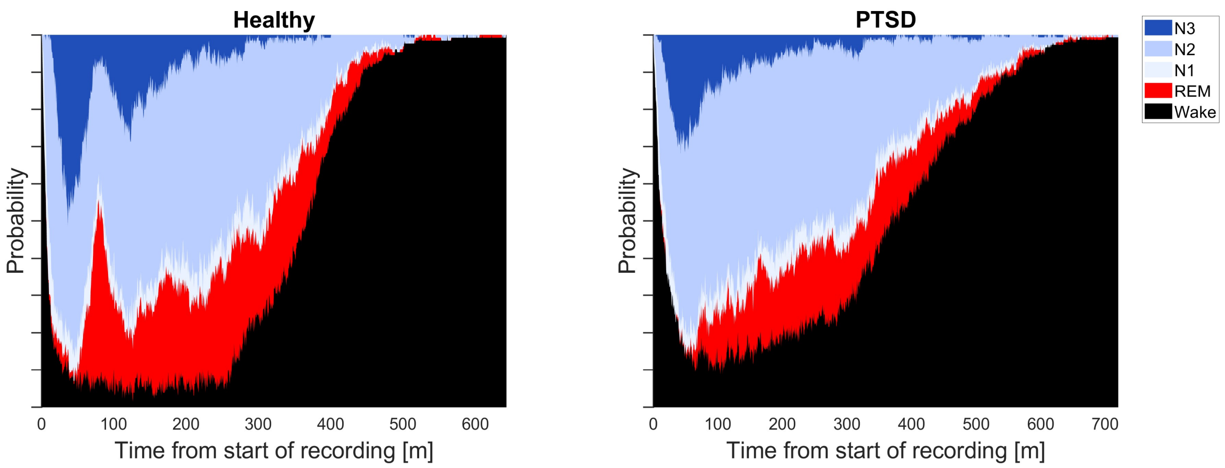


**Figure S2. Time course of sleep stage probability from start of recording.** Stacked area chart depicting all sleep recordings (healthy controls: 130 nights; People with PTSD: 149 nights). The x-axis is aligned to 0, representing the start of recordings. People with PTSD showed a longer wake time before falling asleep (i.e., sleep latency) than healthy controls. PTSD, post-traumatic stress disorder; REM, rapid eye movement; WASO, wakefulness after sleep onset.

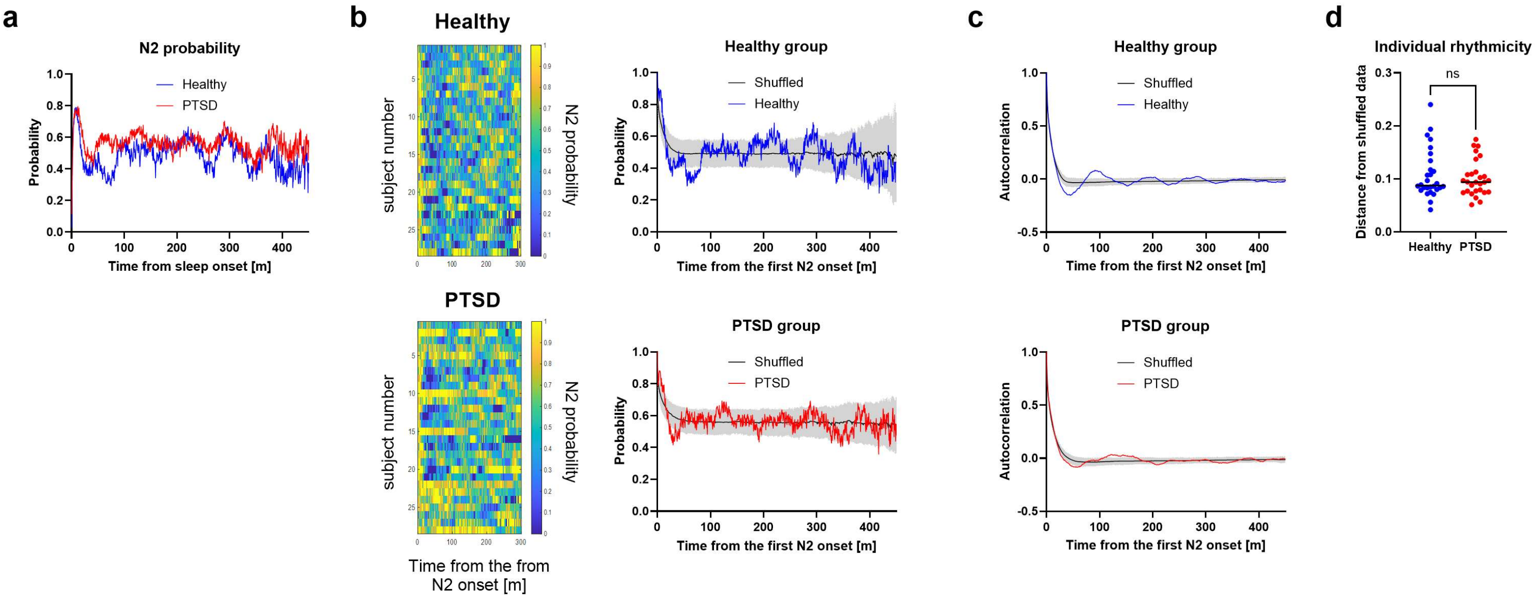


**Figure S3. N2 rhythmicity.** (a) Group-average N2 probability calculated from individual averages across multiple nights. The x-axis is aligned to 0, representing the onset of the first sleep episode. Both groups exhibited several small peaks. (b, c) Nights with no N2 or individual average data ending within 203.5 min after the onset of the first N2 were excluded. (b) Pattern of N2 episode occurrence after the first episode. Heatmap showing individual-average probability (b left), group-average probability (b right), and its autocorrelation (c). The x-axes are aligned to 0, representing the onset of the first N2 episode. The shuffled dataset consists of 1000 group averages, each derived from individual averages across multiple nights, with N2 episodes and their intervals shuffled within nights. The black line is the mean of the shuffled data, and the shaded areas represent the 95% CI derived from shuffled dataset distributions. People with PTSD exhibited few significant peaks. (c) Each data point represents the root mean square deviation of individual average autocorrelation from their respective shuffled data, evaluated up to the time point at which ≥50% of the individual recordings remained. People with PTSD and healthy controls showed similar N2 rhythmicity ($p$ = 0.056). CI, confidence interval; ns, not significant; PTSD, post-traumatic stress disorder.

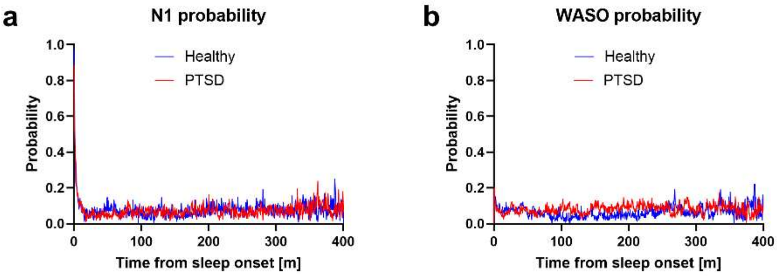


**Figure S4. N1 and WASO rhythmicity.** Group-average probability calculated from individual averages across multiple nights. People with PTSD and healthy controls showed no peak. PTSD, post-traumatic stress disorder; WASO, wakefulness after sleep onset.

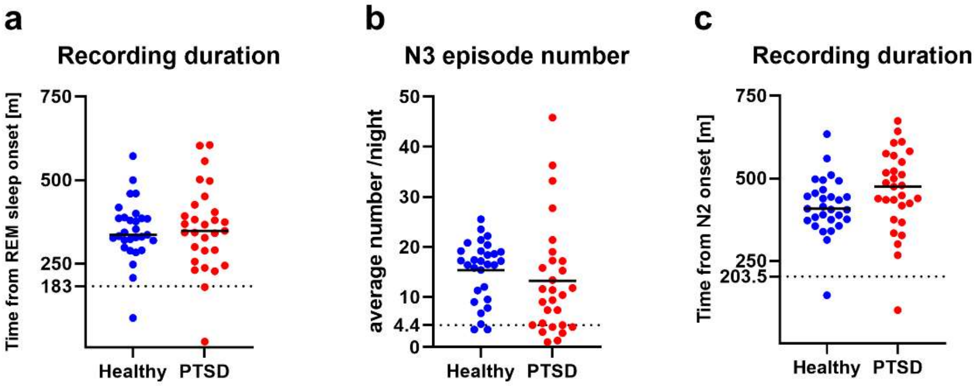


**Figure S5. Data inclusion criteria for rhythmicity analysis.** Nights with no target sleep stages were excluded from REM sleep, N3, and N2 rhythmicity analyses. In addition, some individual average data that did not meet the following criteria were excluded to avoid underestimating the rhythmicity of peaks. (a) Individual average data that ended within 183 min after the onset of the first REM sleep episode (coinciding with the second REM sleep autocorrelation peak in healthy controls) was excluded (healthy controls: n = 1, people with PTSD: n = 2). (b) Individual average data with <4.4 N3 episodes per night on average (corresponding to the first quantile of PTSD group)were excluded (healthy controls: n = 2, people with PTSD: n = 7). (c) Individual average data that ended within 203.5 min after the onset of the first N2 episode (coinciding with the second N2 autocorrelation peak in healthy controls) were excluded (healthy controls: n = 1, people with PTSD: n = 1). PTSD, post-traumatic stress disorder; REM, rapid eye movement.

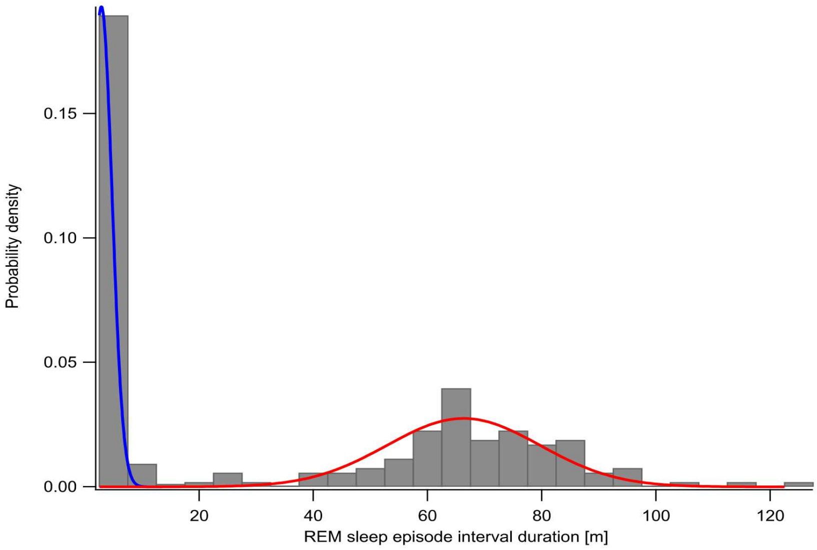


**Figure S6. REM sleep interval distribution with two-Gaussian fit for episode interval analysis.** Distribution of REM sleep episode intervals, fitted with a two-Gaussian mixture model. The histogram represents the probability density of all intervals, and the two fitted Gaussian components are shown as overlaid curves with equal area scaling. The intersection of the two components, located at 17.1 minutes, was used to distinguish between short and long intervals. REM, rapid eye movement.
